# Disrupted functional connectivity of the brain reward system in substance use problems: a meta-analysis of functional neuroimaging studies

**DOI:** 10.1101/2021.12.08.21267481

**Authors:** Jules R. Dugré, Pierre Orban, Stéphane Potvin

**Affiliations:** Research Center of the Institut Universitaire en Santé Mentale de Montréal, Montreal, Canada; Department of Psychiatry and Addictology, Faculty of medicine, University of Montreal, Montreal, Canada

**Author notes:** **Corresponding authors** Jules Roger Dugré, PhD Candidate & Stéphane Potvin, PhD; Research Center of the Institut Universitaire en Santé Mentale de Montréal; 7331 Hochelaga, Montreal, Quebec, Canada; H1N 3V2; /.

## Abstract

**Importance:** Extensive literature suggests that the brain reward system is crucial in understanding the neurobiology of substance use disorders. However, across studies on substance use problems, evidence of reliable disruptions in functional connectivity is limited.

**Objective:** To uncover deficient functional connectivity with the brain reward system that are reliably associated with substance use problems, by meta-analytically synthesizing results of functional brain connectivity studies on substance use problems.

**Data Sources:** Identification of relevant functional brain connectivity studies on substance misuse was done using PubMed, Google Scholar and EMBASE (until September 2021) with the following terms: cannabis, cocaine, substance, methamphetamine, amphetamine, alcohol, tobacco, nicotine, functional connectivity, resting-state, task-based connectivity, psychophysiological interaction.

**Study Selection:** Guidelines of the Preferred Reporting Items for Systematic Reviews and Meta-analyses were followed, Publications were included if they reported stereotactic coordinates of functional brain connectivity results on individuals with substance use problems without a comorbid major mental illness or organic impairment.

**Data Extraction and Synthesis:** Spatially convergent brain regions across functional connectivity studies on subjects with substance use problems were analyzed using Activation Likelihood Estimation meta-analysis.

Altered connectivity with regions of the brain reward system was performed carried out through voxelwise seed-based meta-analyses. Subanalyses were performed to examine mediating factors such as severity of illness, connectivity modalities and types of substances.

**Main Outcomes and Measures:** Identification of deficits in functional brain connectivity with the reward system across studies on substance use problems.

**Results:** Ninety-six studies using a seed-based connectivity approach were included, representing 5757 subjects with substance use problems. In subjects with substance use problems, the ventromedial prefrontal cortex exhibited hyperconnectivity with the ventral striatum, and hypoconnectivity with the amygdala and hippocampus. Executive striatum showed hyperconnectivity with motor thalamus and dorsolateral prefrontal cortex, and hypoconnectivity with anterior cingulate cortex and anterior insula. Finally, the limbic striatum was found to be hyperconnected to the orbitofrontal cortex, and hypoconnected to the precuneus, compared to healthy subjects.

**Conclusions and Relevance:** The current study provided meta-analytical evidence of deficient functional connectivity between brain regions of the reward system and cortico-striato-thalamocortical loops in addiction, in line with current influential neurobiological models. These results are consistent with deficits in motivation and habit formation occurring in addiction, and they also highlight alterations in brain regions involved in socio-emotional processing and attention salience.

**KEY POINTS:** *Question:* What functional brain connectivities with the brain reward system are reliably disrupted across studies on substance use problems?

*Findings:* Subjects with substance use problems exhibited deficient connectivity between the ventromedial prefrontal cortex and subcortical structures including the ventral striatum, amygdala, and hippocampus. Executive striatum showed hyperconnectivity with motor thalamus and dorsolateral prefrontal cortex, and hypoconnectivity with anterior cingulate cortex and anterior insula. Altered connectivity between limbic striatum and core regions of the default mode network was also observed.

*Meaning:* Deficient functional brain connectivity along the cortico-striato-thalamocortical loops may reflect deficits in habit formation, socio-emotional and salience processing in addiction.

## 1. INTRODUCTION

According to the National Epidemiologic Survey on Alcohol and Related Conditions^1^, approximatively 3.9% individuals in the United States have exhibited at least 2 symptoms of a substance use disorder (SUD) in the last year, and this rate increases up to 9.9% when considering the lifetime prevalence. Problematic substance use is often associated with a wide range of impairments, including elevated risk for suicidality and incarceration, homelessness, psychological and health problems, which represent substantial costs to society^2-4^. Despite the high prevalence rate of SUDs in general population, the neurobiological processes involved in SUDs are only partially understood and evidence demonstrating shared and specific neurobiological markers between substances remains limited.

Preclinical research across several decades (using self-stimulation and self-administration paradigms), has consistently shown that the dopaminergic mesocorticolimbic system plays a key role in both short- and long-term effects of most psychoactives substances^5,6^. Human neuroimaging research has also highlighted similar findings. Indeed, several positron emission tomography (PET) studies have revealed that the *acute* administration of most psychoactive substances (e.g., alcohol, nicotine, stimulants) increases synaptic dopamine levels in the ventral striatum^7^ (VS), thereby producing their euphoric effects. However, after *chronic* administration, neuroadaptations occur which lead to a motivational imbalance whereby drug-associated cues gain motivational value and non-drug rewards become less motivational. For instance, subjects with SUDs may exhibit lower availability of striatal dopamine-D_2_ receptors^7^ and reduced activity in the striatum during the anticipation of (non-drug) rewards^8^. Moreover, several functional magnetic resonance imaging (fMRI) studies on drug cravings have shown that drug cues elicit robust activations of core regions of the brain reward system, including the striatum, perigenual anterior cingulate cortex (ACC) and ventromedial prefrontal cortex (vmPFC)^9-12^.

Meta-analytical evidence from task-based fMRI studies indicates that SUD may also be associated with neural alterations outside the reward pathways. Indeed, given that SUDs are often linked to high impulsiveness and emotion regulation problems^13-16^, recent fMRI meta-analyses indicated substantial neural deficits during cognitive control and emotion processing / regulation tasks. For instance, two recent meta-analyses showed that during cognitive controls tasks, individuals with SUDs display reduced activations in the dorsal ACC (dACC) and dorsolateral PFC^12,17^, which play key roles in executive functions. Moreover, subjects with SUDs display reduced activations in the ventrolateral PFC, ACC, anterior insula and amygdala in response to negative emotional stimuli^15,16^. In line with these findings, studies using structural imaging method found that grey matter deficits in subjects with SUDs are not restricted to regions implicated in the brain reward system (e.g. striatum and vmPFC/mOFC), and are also frequently observed in the anterior insula, thalamus, ACC and the amygdala^18-21^. Finally, in functional brain connectivity studies (both task-based and resting-state modalities), researchers found impaired connectivity between brain regions of the reward system in individuals with SUDs ^22^, with possible alterations in other brain networks such as the frontoparietal and the default-mode networks^23-25^. However, there is currently no meta-analysis that primarily aimed to synthesize results of functional brain connectivity studies, leaving unknown whether the reported patterns of dysconnectivity show adequate reliability across studies.

Through the years, researchers have noticed that neurobiological markers of SUDs may differ between substances. For instance, blunted dopamine activity in the VS is often found in alcohol, nicotine and cocaine use disorder, but less consistently in cannabis use disorder^7^ These results concur with results from the ENIGMA consortium indicating that subjects with alcohol use disorder show grey matter deficits across the brain (prefrontal, temporal, subcortical), whereas individuals with cannabis use disorder only show small differences in cortical thickness compared to healthy controls^21^. Moreover, in fMRI studies investigating drug cravings, it has been shown that cocaine and heroin may produce the most potent activations of the mesocorticolimbic system^9,26^. Additionally, only subtle differences in prefrontal activity are often found between substances^12,17^. Searching for shared and specific neurobiological markers is thus crucial to better characterize substances and their related phenotypes.

In view of the state of evidence, our main objective was to perform a coordinate-based meta-analysis of functional brain dysconnectivity associated with substance use problems. More specifically, analyses were conducted on studies reporting hypo & hyperconnectivity results to investigate brain regions that show consistent dysconnectivity. Considering the prominent role of dopaminergic circuit in SUD^5,6^, a seed-based connectivity meta-analysis was performed using key regions of the brain reward system (e.g. Striatum and vmPFC) as Seeds of Interest (SOI), to examine their potential altered connections across the whole-brain (distant connectivity).

## 2. METHODS

### 2.1. Selection Procedures

#### 2.1.1. Search Strategies

A systematic search strategy, using three search engines (Google Scholar, PubMed and EMBASE), was performed independently by two researchers (JRD & SP) up to December 2020 to identify relevant studies. The following search terms were used: (“*cannabis*” or “*cocaine*” or “*substance*” or “*methamphetamine*” or “*amphetamine* or “*alcohol*” or “*tobacco*” or “*nicotine*”) AND (“*functional connectivity*” or “*resting state” or “task-based connectivity” or “psychophysiological interaction”)*. Additional search was executed by cross-referencing the reference lists of the included articles.

#### 2.1.2. Selection criteria

Flow-chart can be retrieved in Supplementary Figure 1. Articles were included if they met the following criteria: (1) original paper from a peer-reviewed journal, (2) inclusion of individuals with a substance use problem-to-disorder without a comorbid major mental illness or organic impairment, (3) use of resting-state or task-based functional connectivity MRI measure, including region-of-interest (ROI) based mass univariate (ROI-to-ROI) and/or voxel-wise connectivity (Seed-to-Voxels), and/or non-seed-based measures such as regional homogeneity (ReHo), fractional- and amplitude of low-frequency fluctuations (fALFF & ALFF) (local connectivity measures) and voxel-mirrored homotopic connectivity (VMHC) (4) conducted a group comparison or a dimensional analysis measuring brain dysconnectivity associated with severity of substance use). Exclusion criteria were: (1) studies using acute administration approach and (2) intrinsic (e.g., Independent Component Analyses) and effective (e.g., Dynamic Causal Modelling) functional connectivity methodologies. Furthermore, when papers did not report peak coordinates for seed or targets, authors were contacted. Preferred Reporting Items for Systematic Reviews and Meta-Analyses (PRISMA)^27^ and the ten rules for neuroimaging meta-analysis^28^ were followed across the meta-analysis steps.

### 2.2. Statistical Procedure

#### 2.2.1. Activation Likelihood Estimate method

In this meta-analysis, the activation likelihood estimation (ALE) approach (GingerALE version 3.0.2, http://www.brainmap.org/ale/) was employed to examine convergent brain regions across functional connectivity studies, as used recently by our research team^29^. Talairach coordinates were converted into MNI (Montreal Neurologic Institute) space before using them in analyses. Briefly, for each experiment, a modeled activation map (MA) was created by modeling coordinate foci with a spherical Gaussian probability distribution, weighted by the number of subjects in each experiment. This is done to account for spatial uncertainty due to template and between-subject variance ^30^, and ensure that multiple coordinates from a single experiment do not jointly influence the modeled activation value of a single voxel. Voxel-wise ALE scores were then computed as the union of modeled activation maps, which provide a quantitative assessment of convergence between brain activation across experiments. The size of the supra-threshold clusters was compared against a null distribution of cluster sizes derived from artificially created datasets in which foci were shuffled across experiments, but keeping other properties of original experiments (e.g., number of foci, uncertainty). A minimum of 10 experiments per meta-analysis was set, since analyses involving <10 experiments drastically increases the risk that a single experiment drives the results^31^. Finally, we used the following statistical threshold across meta-analyses: p<0.001 at voxel-level and FWE-p<0.05 at a cluster-level with 5000 permutations. Subanalyses were performed using using Chi-Square (χ^2^) and Mann-Whitney U tests to assess whether connectivity results may be driven by age, sex, patients’ status (i.e., SUD versus Users) and fMRI modality (i.e., resting-state versus task-based connectivity). We also assessed, using binomial tests, whether the proportion of a given type of substance in a significant cluster was statistically different than its base rate (e.g., proportions of studies on the given substance in the meta-analysis).

#### 2.2.2. Seed-Based Dysconnectivity

Connectivity pattern of a given brain region was investigated using seed-based ALE meta-analysis. Analyses specifically aimed to examine voxels regions that are reliably dysconnected to a particular region (Seed-of-interest, SOI)^29^. Given the importance of the Reward system (e.g. Striatum and vmPFC/OFC) in SUDs^8,32^, we used the 3-subdivisions of the Striatum (i.e. Limbic, Executive and Sensorimotor Striatum from the Oxford-GSK-Imanova Striatal Connectivity Atlas) as well as the vmPFC/OFC (Frontal medial orbital, Rectus and mOFC, Automated anatomical labelling atlas ^33^) mask images (See Supplementary Material). First, we extracted experiments reporting a seed coordinate that fell within the SOI image. ALE meta-analyses were then run on these experiments for each of the SOI. Significant results therefore suggest that clusters are *functionally connected* with their respective SOI across experiments. Meta-analyses were done separately for hyper (SU>HC) and hypo-connectivity (HC>SU) since ALE method does not inherently use effect sizes. Although it would have been relevant to perform analyses using other SOIs, such as brain regions involved in executive functions (e.g., dorsolateral prefrontal cortex & dorsal anterior cingulate), these brain regions were used as SOIs in less than 10 experiments. Thus, they were left out from the meta-analysis.

### 2.2.3. *Non-Seed-Based Connectivit*y

Non-SBC studies included regional homogeneity, fractional- and amplitude of low-frequency fluctuations (local connectivity measures) and voxel-mirrored homotopic connectivity. Studies using large-scale networks analyses (e.g., independent component analysis) were only added in the NSBC meta-analysis if they reported voxel-wise group effects within specific networks (e.g., within-DMN), since these studies report significant peak coordinates of dysconnected voxels.

## 3. RESULTS

### 3.1. Included Studies in Seed-Based Connectivity

A total of 96 studies were included in the meta-analysis on seed-based connectivity (See Supplementary Material). Seed-based studies comprised a total of 5757 subjects, with samples’ average of 33.18 years old (SD=9.29) and 74.85% of males (range: 25-100). Moreover, they contained 59 studies that included SUD patients and 37 studies on substance Users. Distribution of substance categories was: 23 studies on Alcohol, 18 studies on Cannabis, 18 studies on Nicotine, 30 studies on Stimulants (i.e., Cocaine, Amphetamines and Methamphetamines), 4 studies with polysubstance and 3 with other substance (e.g., Heroine).

#### 3.1.1. Orbitofrontal & Ventromedial prefrontal cortex

Using the OFC/vmPFC as a SOI, experiments were extracted for hyperconnectivity (k=13, 21 foci, 2696 subjects) and hypoconnectivity (k=15, 27 foci, 2515 subjects). Results showed that the OFC/vmPFC was hyperconnected to the left ventral striatum and hypoconnected to the right hippocampus and amygdala (Table 1, Figure 1). Subanalyses revealed that the OFC-vSTR connectivity was associated with more samples of SUD patients compared to samples of users (X2=4.17, p<0.041). No other subanalyses yielded significant findings for dysconnectivities with the vSTR, the hippocampus and the amygdala (Ps > 0.218).

**Table 1.**
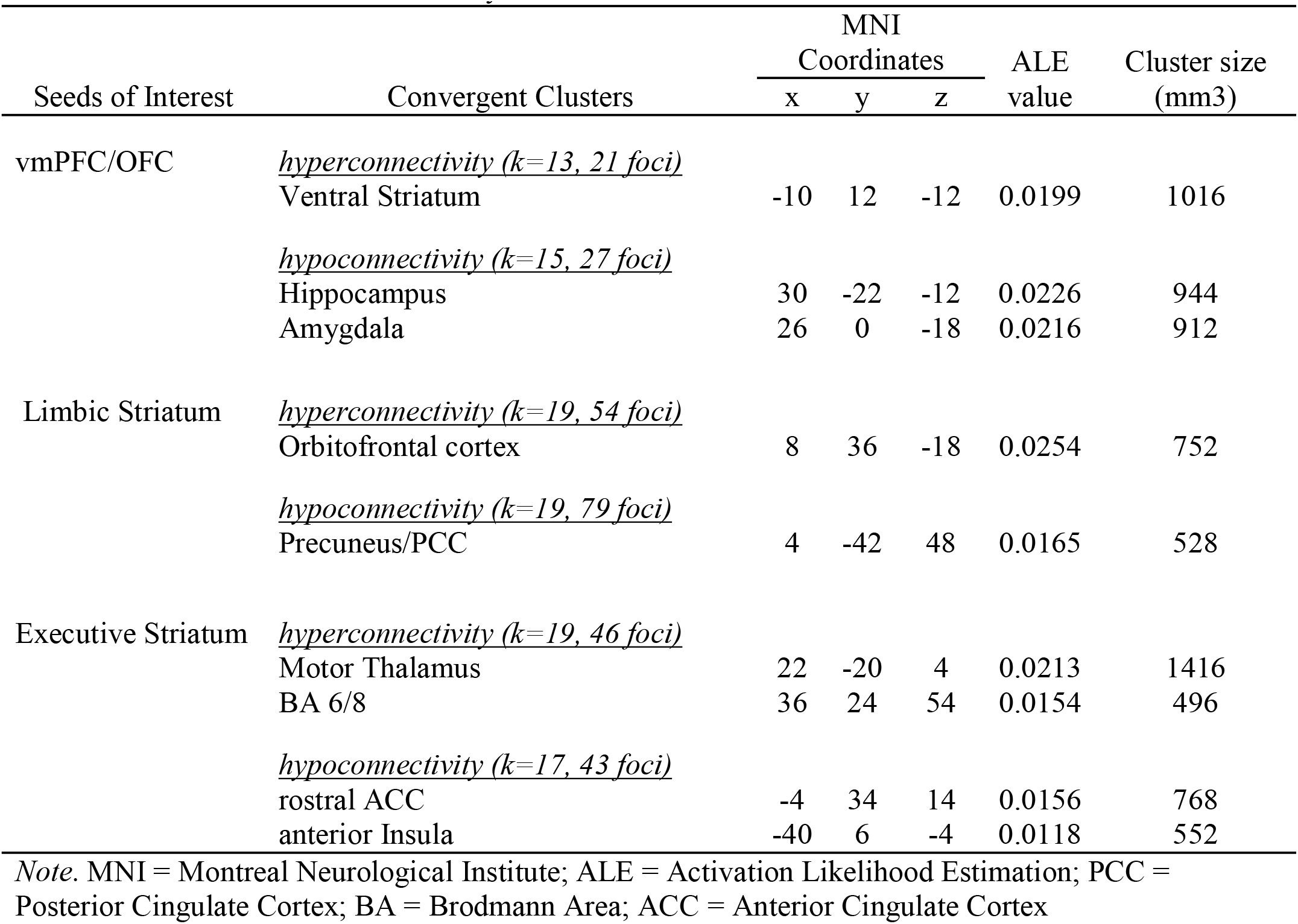
Results of the ALE meta-analyses on Seeds of Interest

**Figure 1.**
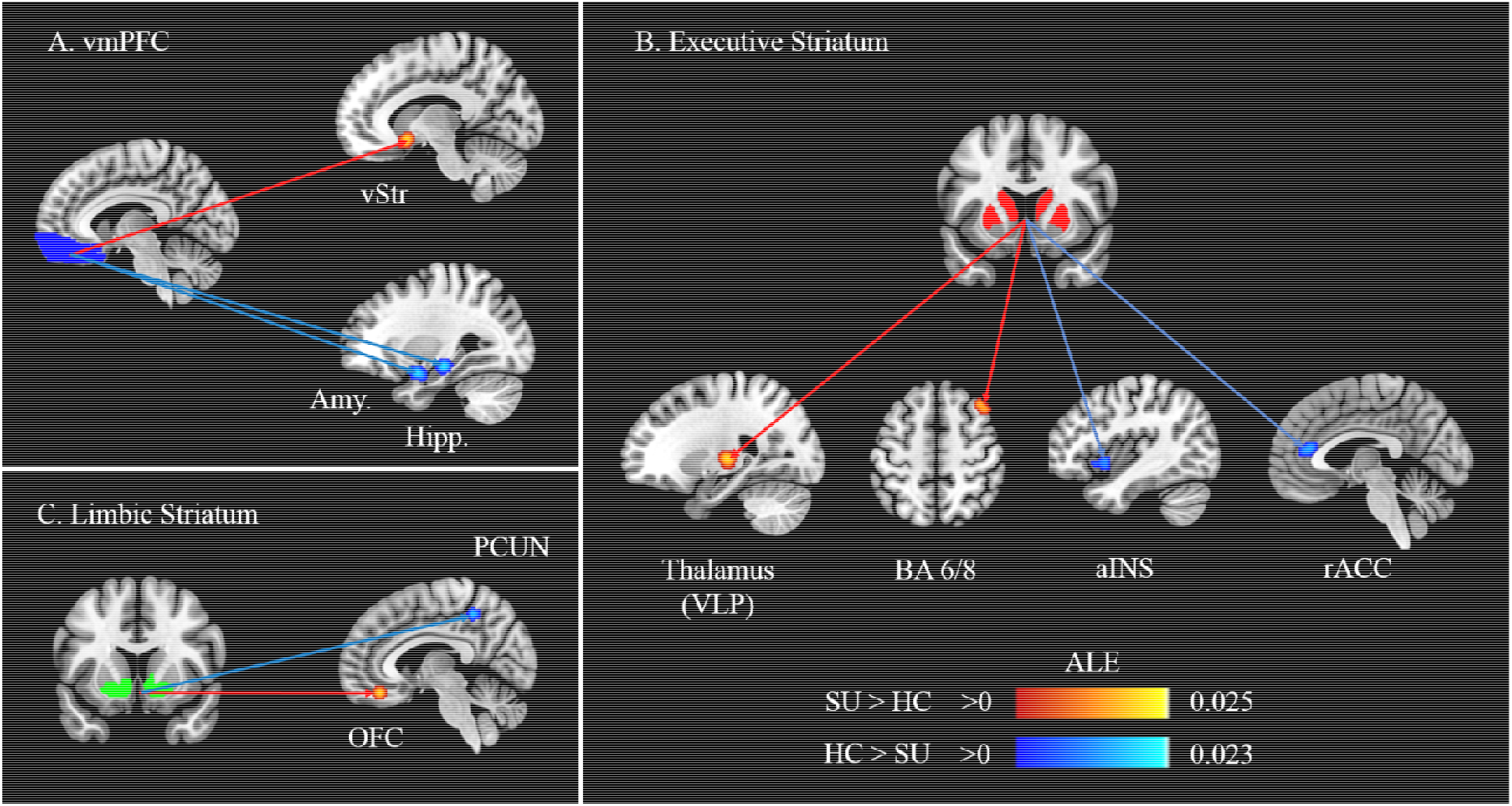
Summary of results from the ALE meta-analyses. A. Meta-analysis on disrupted connectivity associated with the ventromedial Prefrontal Cortex (Seed-of-Interest), B. Meta-analysis on disrupted connectivity associated with the Executive Striatum (Seed-of-Interest) and C. Meta-analysis on disrupted connectivity associated with the Limbic Striatum (Seed-of-Interest). Coloured lines represent hyperconnectivity (Red) and hypoconnectivity (Blue) in individuals with substance use / problems. vmPFC = ventromedial prefrontal cortex; vStr = ventral Striatum; Amy. = Amygdala; Hipp. = Hippocampus; PCUN = Precuneus; OFC = Orbitofrontal Cortex; VLP = Ventrolateral posterior; BA = Brodmann Area; aINS = anterior Insula; rACC = rostral Anterior Cingulate Cortex.

#### 3.1.2. Striatal subregions

Striatal subregions, defined by the Oxford-GSK-Imanova Striatal Connectivity Atlas, were used as SOIs. Limbic Striatum was used as a seed in 19 studies reporting hyperconnectivity (54 foci, 1046 subjects) and in 19 studies reporting hypoconnectivity results (79 foci, 1193 subjects). Meta-analysis revealed that the limbic striatum was hyperconnected to the OFC, and hypoconnected to the Precuneus/PCC (Table 1, Figure 1). No subanalysis yielded significant results for both the OFC and the Precuneus/PCC (Ps > 0.109). Binomial tests revealed that Nicotine was marginally overrepresented compared to its base rate in Limbic Striatum-OFC connectivity (p=0.067), whereas no other effect was found for other drugs nor for the precuneus/PCC (Ps>.171).

Furthermore, the Executive Striatum was found to be hyperconnected (k=19, 46 foci, 976 subjects) to the Motor Thalamus (ventral lateral posterior nucleus) and the Brodmann area 6/8, and hypoconnected (k=17, 43 foci, 638 subjects) to the rostral ACC and anterior insula (Table 1, Figure 1). Subanalyses yielded no significant findings for these connections (Ps < 0.119).

Regarding the Sensorimotor Striatum, no meta-analysis was run due to the small number of experiments for both hyperconnectivity (k=3) and hypoconnectivity (k=5).

### 3.2. Non-Seed-Based Dysconnectivity

A total of 31 experiments used a NSB to measures dysconnectivity in SU patients (total of 882 patients). Samples’ average age was 35.07 years old (SD=10.74) and 84.57% of participants were males (range: 0-100). A total of 9 studies focused on Alcohol, 5 on Cannabis, 6 on Nicotine, 9 on Stimulants, 1 on polysubstances and 1 on heroin. However, analyses were not performed due to the large heterogeneity in measures of NSB connectivity (see Supplementary Material) and the lack of a sufficient number of experiments per NSB measure.

## 4. DISCUSSION

To our knowledge, this is the first meta-analysis that aims to characterize deficits in functional brain connectivity associated with substance use problems. Using 96 studies (5757 subjects) that adopted a seed-based approach, we observed that brain regions involved in the reward system were disconnected with several brain structures crucial to our understanding of substance use problems. For instance, meta-analysis on vmPFC/OFC revealed hyperconnectivity with the ventral striatum and hypoconnectivity with the hippocampus and amygdala. Furthermore, the limbic striatum showed hyperconnectivity with the orbitofrontal cortex and hypoconnectivity with the precuneus/PCC, whereas the executive striatum was hyperconnected to the motor thalamus and BA 6/8 and hypoconnected to the rostral ACC and anterior insula. These results provide substantial insight into our understanding of the neural mechanism underlying substance use problems.

Striatal deficiencies have directed neurobiological research on substance use for the past decades. Here we showed that subjects with substance use problems showed impaired striatal connectivity with limbic and executive subregions. In healthy subjects, the VS (limbic striatum) show strong connectivity with the vmPFC/OFC ^34,35^. Indeed, past researches have robustly shown that these regions are involved in reward prediction and processing of the magnitude of received reward ^36,37^, defining both regions as crucial nodes of the reward system^38^. In our study, we observed that SUD samples (compared to users) were more likely to show VS-vmPFC/OFC dysconnectivity. These are consistent with results of task-based fMRI studies on subjects with substance use problems showing reliable activations of brain reward regions in response to drug cues ^9,39,40^. Taken together, these results are in line with influential neurobiological models which state that addiction is characterized by an acquired over-valuation of the motivational value of drug stimuli, stemming from the neural sensitization of dopamine-related reward systems^41,42^. In addition, we observed that substance use problems were associated with a reduced connectivity between the VS and the Precuneus/PCC, which is a core region of the default-mode network and plays a well-established role in self-referential processes^43,44^. Interestingly, the functional connectivity between the VS and the precuneus/PCC has also been found in other forms of addiction such as internet gaming disorder^45^. Such results are consistent with newer neurobiological models emphasizing that importance of impaired self-awareness in human addiction^46^.

In healthy subjects, the dorsal part of the striatum (executive) is mainly connected to the aMCC/pre-SMA, insula, thalamus and dlPFC^34,47-49^. In line with these results, we found that the executive striatum was hyperconnected with the motor thalamus and the dlPFC. Moreover, we observed that the executive striatum was hypoconnected with the rACC and anterior insula, which are the two core regions of the salience network^44^. Past systematic reviews and meta-analyses on functional neuroimaging studies reported that SUD subjects showed prominent hypoactivity in these particular regions during cognitive control tasks^12,50,51^, but hyperactivity in responses to drug cues^12,50^. As theorized by Everitt and Robbins^52^, repeated use of substances may both exaggerate the attention salience to drug cues (e.g., ACC and anterior insula) and alter executive control (e.g., dlPFC, motor thalamus) over such stimuli. Future research may seek to examine the specific role of these dysconnectivities in relation to SUD symptoms.

Meta-analytical evidence of fMRI studies in healthy subjects indicates a crucial role of the vmPFC in subjective valuation and emotional decision-making^53^, whereas vmPFC lesion studies found increased risk appetite under hot decision conditions ^54-56^. In our meta-analysis, we not only observed a hyperconnectivity between the vmPFC/OFC and VS, but also a hypoconnectivity between the former region and the hippocampus/ amygdala in subjects with substance use problems. One possible interpretation is that the vmPFC may compute subjective valuation by weighting the anticipated cost (amygdala) and benefits (VS), in order to adapt subsequent behaviors ^57^. Hence, a reduced vmPFC-Amygdala connectivity in SU subjects would bias the cost–benefit analysis, resulting in a systematic over-estimation of benefits (vmPFC-VS) over the costs (vmPFC-Amygdala). This concurs with the fact that SU subjects compulsively use and seek drugs despite major negative consequences (e.g., neglecting health, work, relationships).

Limitations of the current meta-analysis need to be acknowledged. First, to achieve 80% power to detect effects that are present in approximately 1/3 of the population, a sample size of 17 experiments is required when using cluster-level FWE^31^. Therefore, results from meta-analyses with less than 17 experiments should be interpreted carefully, as it is the case here for the sub-analyses performed on specific substances. Second, regarding the meta-analyses on subtypes of substance, differences in seed selection between substances may have confounded our results. Thus, larger sample sizes and direct comparison between subtypes of substances will be needed to truly examine the specificity of (or lack of) functional dysconnectivities observed between substance types. Third, most studies in the field have selected seeds from the reward system, which has limited our ability to look at other crucial brain regions. Finally, we performed a coordinate-based meta-analysis rather than a meta-analysis of original statistical brain maps which may produce results with reduced accuracy^58^.

## CONCLUSIONS

By conducting the first meta-analysis of functional connectivity studies in subjects with substance use problems, we observed that the vmPFC/OFC and striatal subregions (Limbic and Executive Striatum) showed distinct dysconnections with brain regions involved in emotion processing, self-valuation, habit formation and attention salience. These findings concur with past neurobiological models of addiction highlighting the crucial role of both the VS and DS in maladaptive drug-seeking behaviors. Future studies should investigate these dysconnections in relationships with more precise phenotypes such as age-of-onset, severity of SUDs and withdrawal. Longitudinal studies are required to better understand how the neural alterations reported here emerge and evolve during the different phases of addiction. Finally, additional whole-brain functional connectivity studies are required to determine if other brain regions / neural networks play a key role in the neurobiology of addiction.

## Data Availability

All data produced in the present study are available upon reasonable request to the authors

